# Local protection bubbles: an interpretation of the decrease in the velocity of coronavirus’s spread in the city of São Paulo

**DOI:** 10.1101/2020.08.11.20173039

**Authors:** José Paulo Guedes Pinto, Patrícia Camargo Magalhães, Gerusa Maria Figueiredo, Domingos Alves, Diana Maritza Segura-Angel

## Abstract

After four months of dealing with the pandemic, the city of São Paulo entered a phase of relaxed social-distancing measures in July 2020, and saw its social isolation rate fall at the same time as the number of cases, deaths, and hospital bed occupation declined. We use a calibrated multi-agent model to describe these dynamics. We assert here that this phenomenon can be understood as the result of local protective bubbles formed in the city’s sub-environments at the same time that there was an exhaustion of contagion networks. Both reduce the velocity of the virus’s spread, causing temporary reductions in the epidemic curve, albeit in an unstable equilibrium. These local bubbles can burst anytime and anywhere due to the reintroduction of a few infected people at the same time that there is a reduction in non-pharmaceutical interventions (NPI), such as social-distancing practices. It is important to stress that this hypothesis aligns with the dynamics of the virus’s spread observed so far, without needing ad hoc suppositions about natural collective immunity thresholds or heterogeneity in the population’s transmission rate, which come with the risk of making mistaken predictions that may could lead to the loss of many lives. The safe way to move ahead is to continue doing all we can to avoid new infections until a vaccine is found that properly and safely creates herd immunity.

## Introduction

Since the first case was reported on February 25 in the city of São Paulo, the biggest city in Brazil and the country’s epicenter for the disease, the spread of the pandemic has demonstrated a complex dynamic in Brazil up to the present moment (July 2020). This is due mainly to the lack of political coordination between federal authorities and local governments, massive inequalities between territories, and the fact that Brazil is a continent-sized country. Brazil is currently on its way to becoming one of the world’s worst cases, second only to the US, with the disease continuing to spread – especially in the more far-flung regions of the country.

Despite this disturbing scenario, many states, municipalities, and even neighborhoods have been pressured to reopen the economy (even before reopening public parks). The state of São Paulo in particular is relaxing the social-isolation measures it put in place (Governo do estado de São Paulo a, 2020; Bazani, 2020) on June 1, when the pandemic was just reaching the cities in the countryside. The outcomes of this policy were essentially as expected: each reopening phase was followed by a deterioration in pandemic statistics. At the end of July, the COVID-19 epidemic in the state of São Paulo had reached its most critical situation (Figure 1-top).

But there was one interesting exception: the city of São Paulo. After entering a more relaxed stage of social isolation in July, the city saw its social isolation rate fall along with the number of new cases, deaths, and hospital beds occupied, as shown in Figure 1 (bottom).

**Figure 1:**
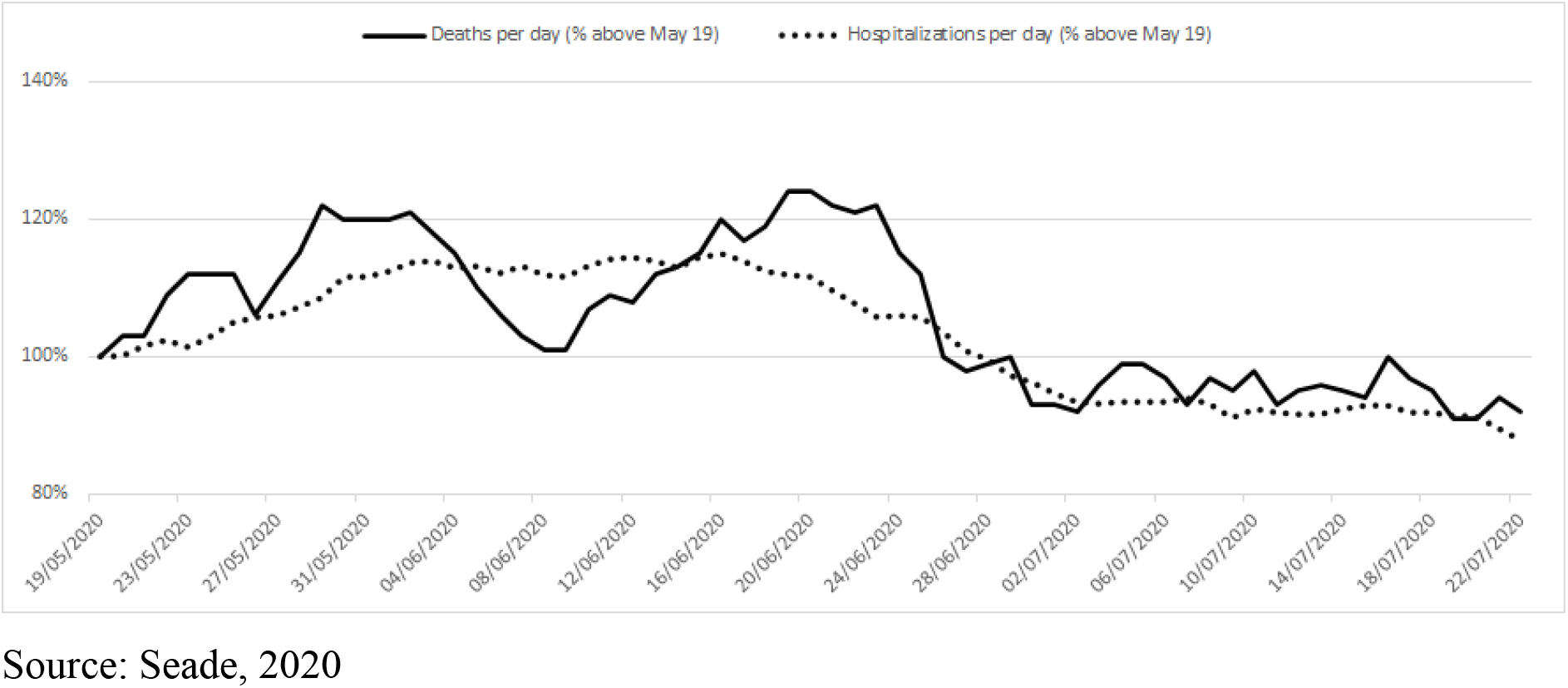
Percentage of deaths (continuous line), hospitalization (dotted line) per day for the state of São Paulo (top) and the city of São Paulo (bottom). The data shows the 7-day moving average, normalized for May 19.

Along with these phenomena, two seroprevalence surveys were published for the city of São Paulo that showed a maximum of 12% of the population being immune at the end of June (SoroEpi MSP, 2020).

This information launched a public debate that tried to diagnose what is causing this effect in the city (Magenta, 2020). There are at least three hypotheses in vogue: the positive outcome of non-pharmaceutical interventions (NPI), herd immunity, and protective bubbles accompanied by the exhaustion of the social-contagion network.

According to the first explanation, non-pharmaceutical interventions carried out by the population – which includes social distancing, washing of hands, and the use of masks – had a positive effect, even if those protocols were only partially adopted and not mandatory (Leung, 2020; Cruz 2020; Greenstone, 2020; Flaxman, 2020; Block, 2020; Candido, 2020; Pike, 2020). But some researchers say that this alone does not explain why the number of hospitalizations has not increased again in the city of São Paulo as social distancing measures were relaxed.

The second explanation suggests that social or herd immunity can be achieved even with low levels of immunity among the population. On the one hand, Grifoni (2020), Gallais (2020), and Sette (2020) analyze studies showing that there might in fact be fewer people susceptible to the coronavirus due to other defenses in the body that can fight the coronavirus – in addition to neutralizing antibodies and T cells, CD4+ and CD8. On the other hand, there are studies (Britton, 2020; Gomes, 2020; Aguas, 2020; Bisin, 2020) that look into heterogeneity within the population, which could lead to a decrease in the percentage of the population that must be infected to achieve herd immunity (leading to a decline in the rate of infection). Using Britton (2020) parameters, the effective herd immunity threshold could be reduced to 43% (or even 34% depending on the scenario), whereas Gomes (2020) shows that this number can drop to 20%. But the authors of the first study explicitly draw attention to the fact that their estimates “should be interpreted as an illustration of how population heterogeneity affects herd immunity, rather than an exact value or even a best estimate.” The limits of this study were even emphasized by Britton (2020).

Some other issues should be considered in the herd immunity explanation. Although SARS-CoV-2 specific T cells are present in individuals with no history of this infection (Grifoni, 2020; Braun, 2020) both the effect and duration of pre-existing memory of SARS-CoV-2 are little known (Le Bert, 2020). In addition, immunity surveys of various territories, such as Spain (5% Pollán, 2020), Wuhan (3.2-3.4%, Xu, 2020), Geneva (10-8%, Strii, 2020), and São Paulo (9.2-13%, SoroEpi MSP, 2020), indicate that less than 30% of the population was infected when the curves for both infections and deaths began to decrease.

The third explanation – our hypothesis – will be developed throughout this article. In short, we do not need to assume collective immunity (or fewer people being susceptible to coronavirus), nor ignore the fact that people are easing their social-distancing practices. We argue that this phenomenon is due to local bubbles of protection that emerge in the city’s sub-environments in the absence of contagion networks. They reduce the velocity of the virus’s spread, causing short and temporary reductions in the epidemic curve – but manifest as an unstable equilibrium.

## Methodology

To simulate the coronavirus’s dispersion in the environment we used “MD corona,” a model developed by us (Guedes Pinto, 2020) which is based on complex multi-agent variables. Unlike (SIR-based) multiple-differential equations and statistical-model approaches, the multi-agent model (Ajelli, 2010; Venkatramanan, 2017) doesn’t necessarily rely on pandemic data (cases, deaths, and recoveries, for instance) to make predictions about the epidemic curve. The phenomenon is described by the successive interactions between individual subjects with particular properties and actions (Wilensky, 2009).

In our multi-agent model, a number of people (agents) are aleatorily placed onto a grid, and they move aleatorily (up/down/left/right). The dynamics of virus transmission will depend on whether the interaction between two or more agents (infected, immune, or susceptible to infection by the virus) on the grid can result in one agent infecting the others. The parameters that directly affect the epidemic outbreaks are the virus-transmission period (eighteen days, WHO, 2020), the immunity period (one year, to avoid the interference of this parameter in our simulations), the initial number of infected individuals (one agent in the grid), the effective transmission probability, the number of agents in the environment, and the rate of social distancing practiced by the population.

For this study, the first three parameters listed above were fixed in our methodology (Magalhaes, 2020), whereas the final three are explained as follows.

The social-distancing rate practiced by the population is a dynamic parameter in the model that slows down the spread of the virus by not allowing a portion of the population to move, and it can vary while the simulation is running to describe changes in social distancing over time.

The effective transmission probability (varying from zero to a hundred percent) is directly related to an index – which can be either the Human Development Index (HDI) or the Coronavirus Protection Index (CPI) developed by Ação Covid-19a (2020), with the latter being based on the Surroundings Index (S.I.) methodology (Ranieri, 2016). The CPI index takes into account the characteristics of the health system, human development, and territorial indicators, which is a more complete and appropriate index to describe the vulnerability of a given territory to the coronavirus transmission when compared with HDI. Both HDI and CPI are broken down into five levels: very high, high, medium, low, and very low (see Figure 2 for the HDI and CPI), which measures the vulnerability of the neighborhood or city.

**Figure 2.**
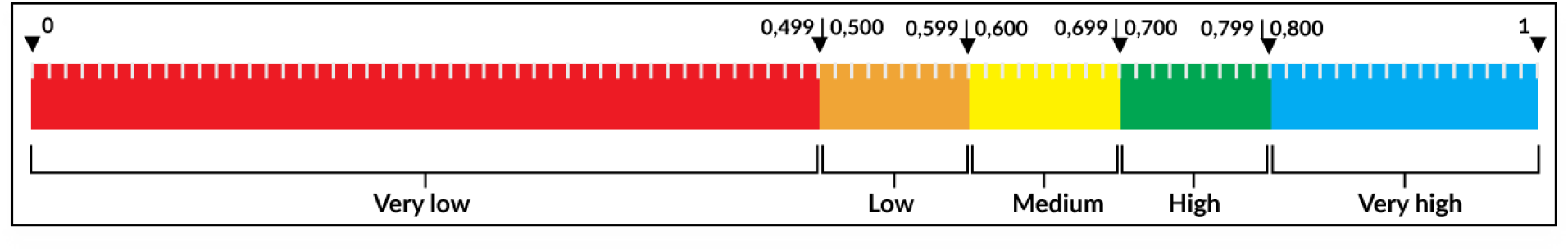
Ranges of HDI/CPI

The connection between the levels shown above and the transmission probabilities, as well as the number of agents in the environment, is fixed by a calibration procedure.

The model can be calibrated by connecting the number of agents in the grid to population density and the effective transmission probability to the respective grade of HDI/CPI. In the case of the city of São Paulo, the model was calibrated by comparing the number of agents in the grid, with its 8,054.7 inhab./km^2^ population density and the effective transmission probability, to the respective scale of CPI (0.79) for São Paulo, which is high. Applying the known history of social confinement for São Paulo (Governo do estado de São Paulo b, 2020) as shown in Table 1 of Appendix A, we fixed the effective transmission probability at 40% in the simulator to match the results of the seroprevalence surveys for 369 agents in the grid.

As for all the multi-agent models, “MD Corona” is a stochastic process that depends on the initial conditions randomly determined by the simulator each time (the relative position of infected and immobilized individuals). To account for this fluctuation we ran all the simulated scenarios 100 times with the aid of a Python program and examined the average results. A more complete stochastic analysis could investigate the different trends in the simulation results, identifying the behavioral change threshold positions, i.e., where the curve’s inflections are. This will be done in an upcoming publication.

In Figure 3, we show that, after calibrating, the simulation using the history of social distancing in the city (Table 1, Appendix A) yields 9.38% of the population infected on average (the colored curves represent each simulation) and 0.08% mortality from Covid-19 on average (or 0.75% of the infected total) on June 22, 118 days after the first reported case, which is compatible with the seroprevalence (immunity) surveys.

**Figure 3.**
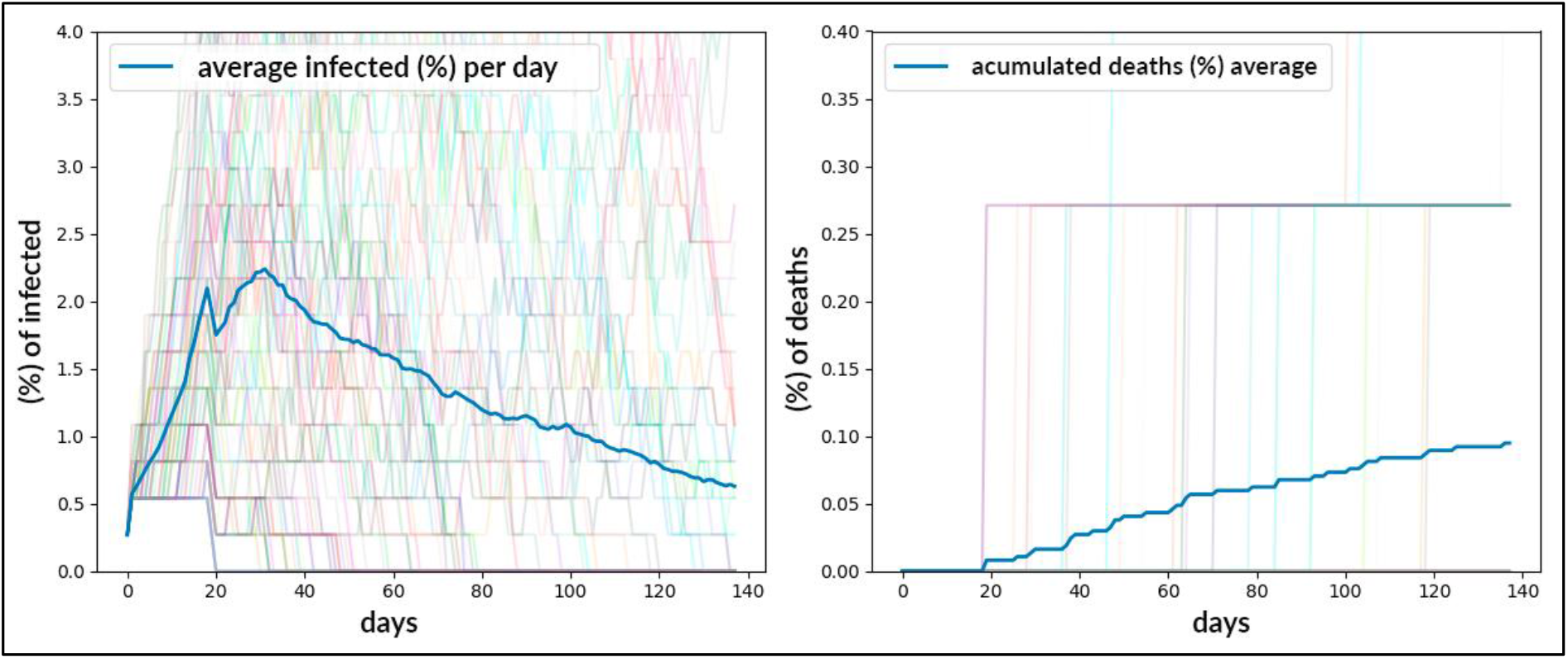
Dispersion dynamics for the coronavirus in the city of São Paulo through July 12, 2020, calibrated on June 22.

## Results

Using the calibrated model, we can make predictions about the epidemic curve and explore different scenarios. The multi-agent simulator’s dynamics describe a closed system, with interactions only between agents in the same local environment. However, we know that as a result of reduced social-distancing measures there will be increased interaction between individuals from different neighborhoods or cities (or even countries), which could reintroduce the virus in that given environment. To account for this phenomenon, the model considers the possibility of reintroducing infected agents after a certain amount of time.

### Scenario 1: reducing social distancing

In order to investigate the effect of reopening the economy on the evolution of the epidemic curve, in scenario 1 we simulate the situation where social distancing is dramatically reduced to 20%.

In this scenario, we applied the official timeline for the social distancing rate released by the São Paulo state government (described in Table 1 of Appendix A) and added the hypothesis of reducing the social-distancing rate to 20% on average for the next 100 days, starting from July 12. The average epidemic curve after 238 simulation days is shown in Figure 4, and gives us a rate of 12.71% of infected agents and 0.12% of the population dying from Covid-19 (i.e., 0.9% lethality) on the 238th day after the first case of the virus.

**Figure 4.**
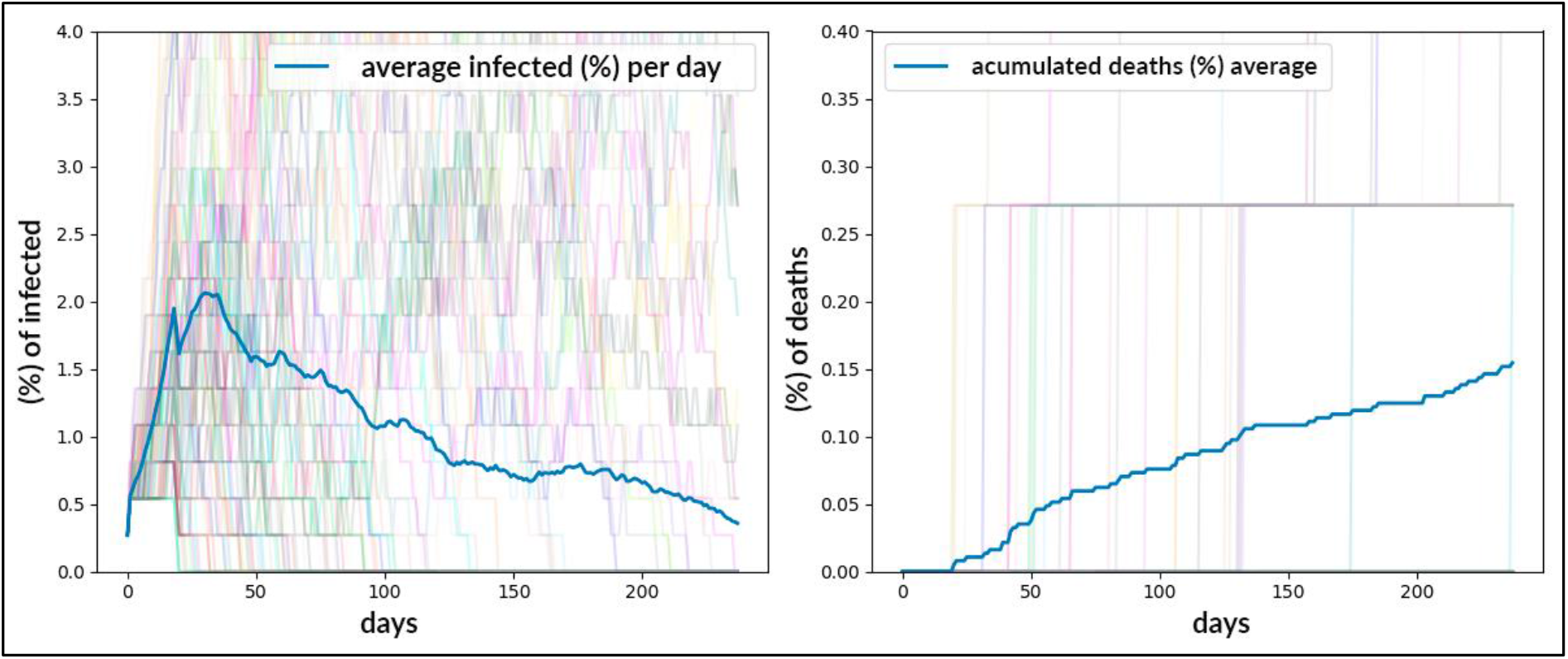
Result of the simulation using MD Corona for scenario 1 and timeline of social distancing for the city of São Paulo, extended by 100 days with a 20% social-distancing rate.

The simulation shows the transmission curve decreasing, even with a drastic reduction in social isolation, with a slight and temporary increase after the 150th day. This effect seems counterintuitive when considering that due to the low immunity rate we are far from reaching herd immunity, and so we developed other hypotheses.

One explanation for this effect can be seen in the model’s simulation grid (Figure 5), where it is possible to observe (circled in yellow) that there are local **bubbles of protection** against the transmission of the coronavirus, with a concentration of infected (red) agents surrounded by immune (gray) ones protecting those agents susceptible to infection (green).

**Figure 5.**
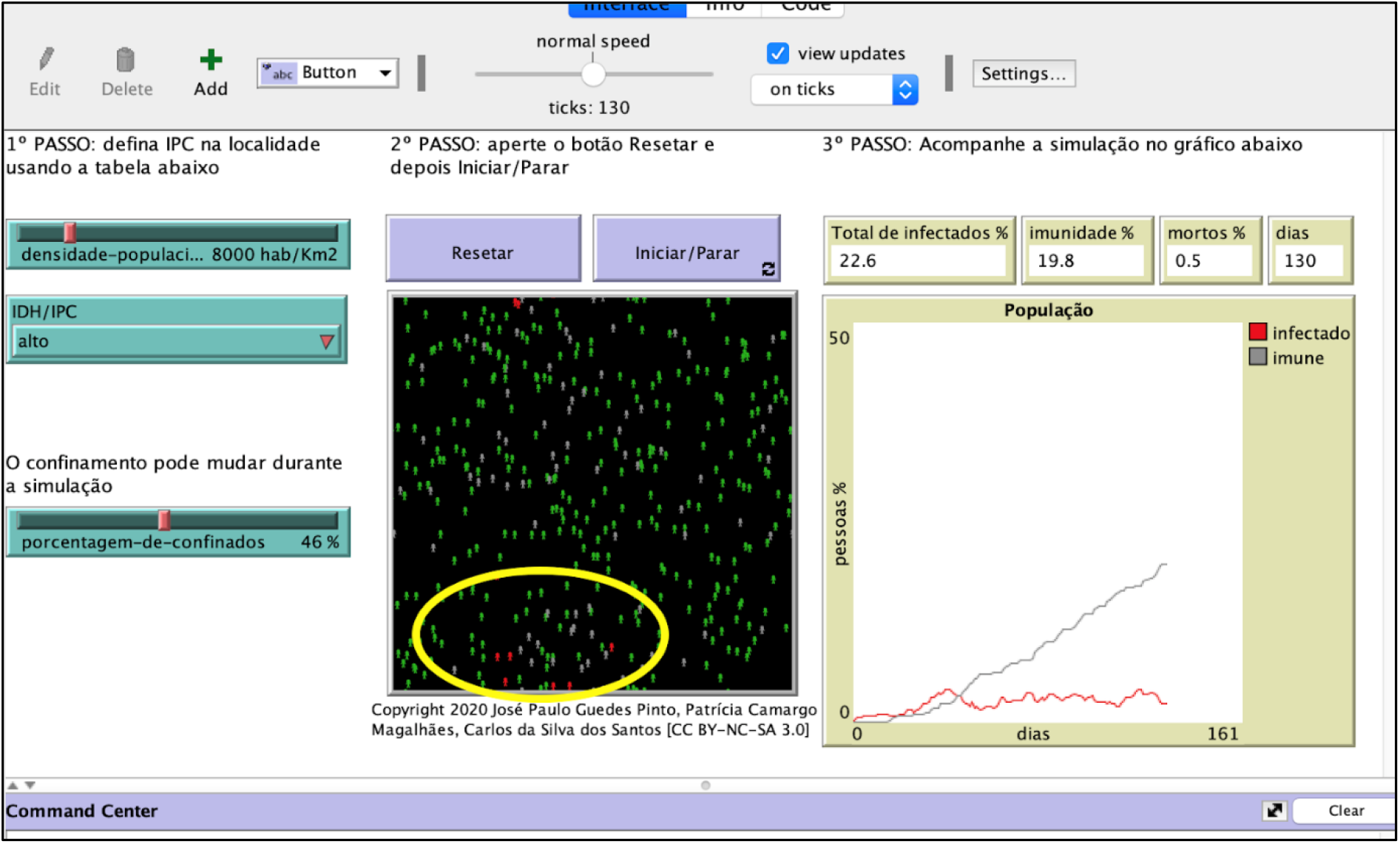
MD Corona simulation on Netlogo software for the city of São Paulo after calibrating for the history of social distancing, provided in Table 1 of Appendix A. The yellow circle highlights the existence of protection bubbles.

Another possible explanation is the **exhaustion of the infection network** in several sub-environments. This occurs when the virus has been present in a portion of the population for a relatively long period of time (138 days since the first case was recorded on February 25, 2020). A substantial number of red (infected) agents carry the virus but don’t find many other green (susceptible) agents in the environment to infect during the transmission period. This is sufficient to decrease the rate of transmission in the given environment.

It is important to emphasize that these two hypotheses, protection bubbles and exhaustion of the network, are local effects. Therefore, they differ from the hypothesis of herd immunity where the virus is supposed to be suppressed in the whole environment, thus achieving a global and stable equilibrium. In our hypotheses, what we have is local equilibrium, possibly unstable, which can be disturbed by the introduction of a new infected agent, which would burst those bubbles and reinitiate the infection networks.

However, we should note that that this dynamic, shown in Figure 5, is exclusively applicable to a density compatible with the city of São Paulo, without taking into account the complex demographic specificities and the Coronavirus Protection Index (CPI) of the different districts within the Municipality.

### Scenarios 2 and 3: reintroducing one sick agent

Working with the situation as it stood on July 12, we then reintroduced one sick agent, representing 0.27% of São Paulo’s population (about 33,000 people) and extended the simulation for 100 days with different social-distancing scenarios. The random reintroduction (within the model’s spatial environment) of one infected agent has the effect of bursting protection bubbles, resulting in a second wave of contamination.

In scenario 2, the social-distancing rate after the reintroduction was set to 20%, and the simulation yields in Figure 6 shows that the second wave may be higher than the first one. This could lead to an increasing risk of collapse in the health system. In this scenario, at the end of 238 simulated days we would have 19.67% of the population infected by the coronavirus, and 0.18% dying from the virus (0.92% lethality).

**Figure 6:**
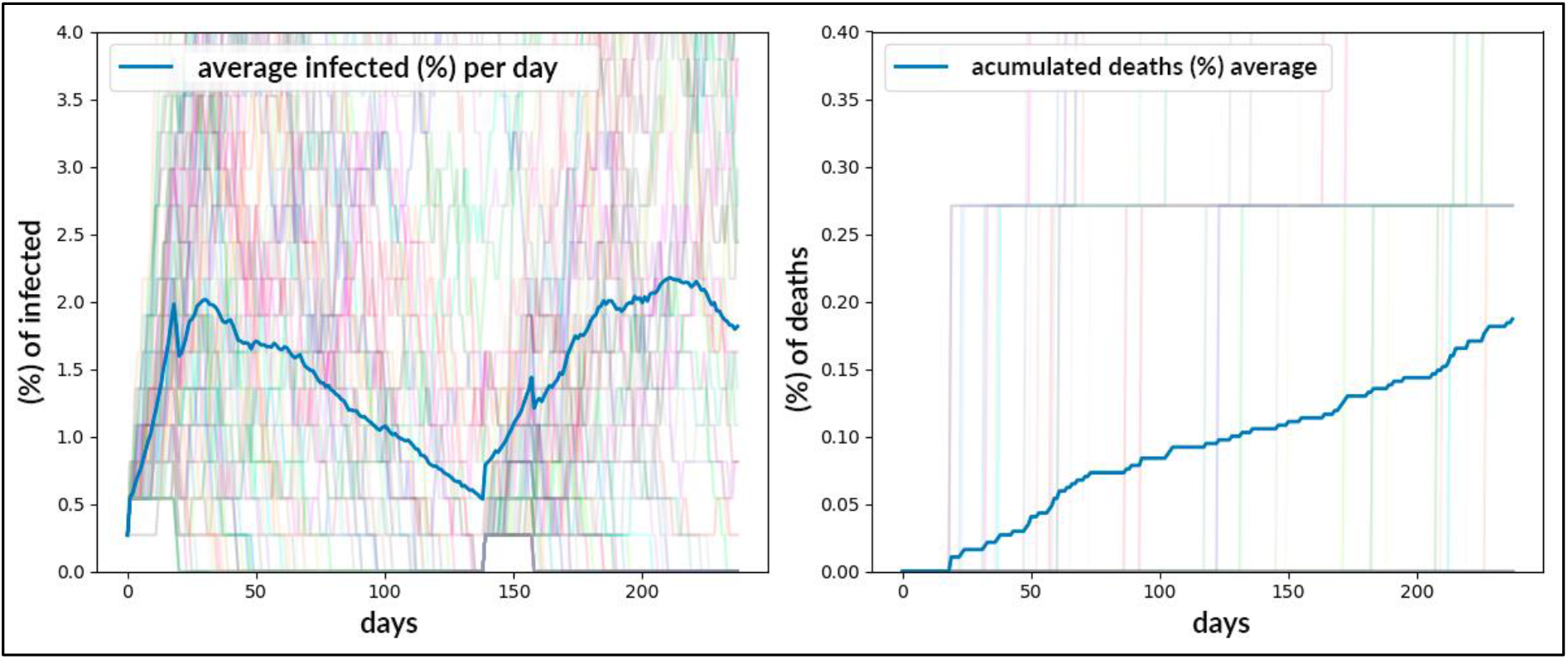
Dynamics of Virus Dispersion in the City of São Paulo following the reintroduction of an infected agent on the 138th day, using the official timeline (Table 1 in Appendix A) of social distancing, extended by 100 days at 20% isolation rate.

In scenario 3, the reintroduction of one infected person on the 138th day of the simulation was accompanied by a 40% social-distancing rate for 100 days. Although we can see in Figure 7 that this also triggers a second wave of a Covid-19 outbreak, because social distancing remains at a higher level than in scenario 2 (40% versus 20%), the second wave is almost always lower than the first one. At the end of 238 simulated days, we would have 15.07% of the population infected by the coronavirus, and 0.13% of the population dying (1.95% of the infected total).

**Figure 7:**
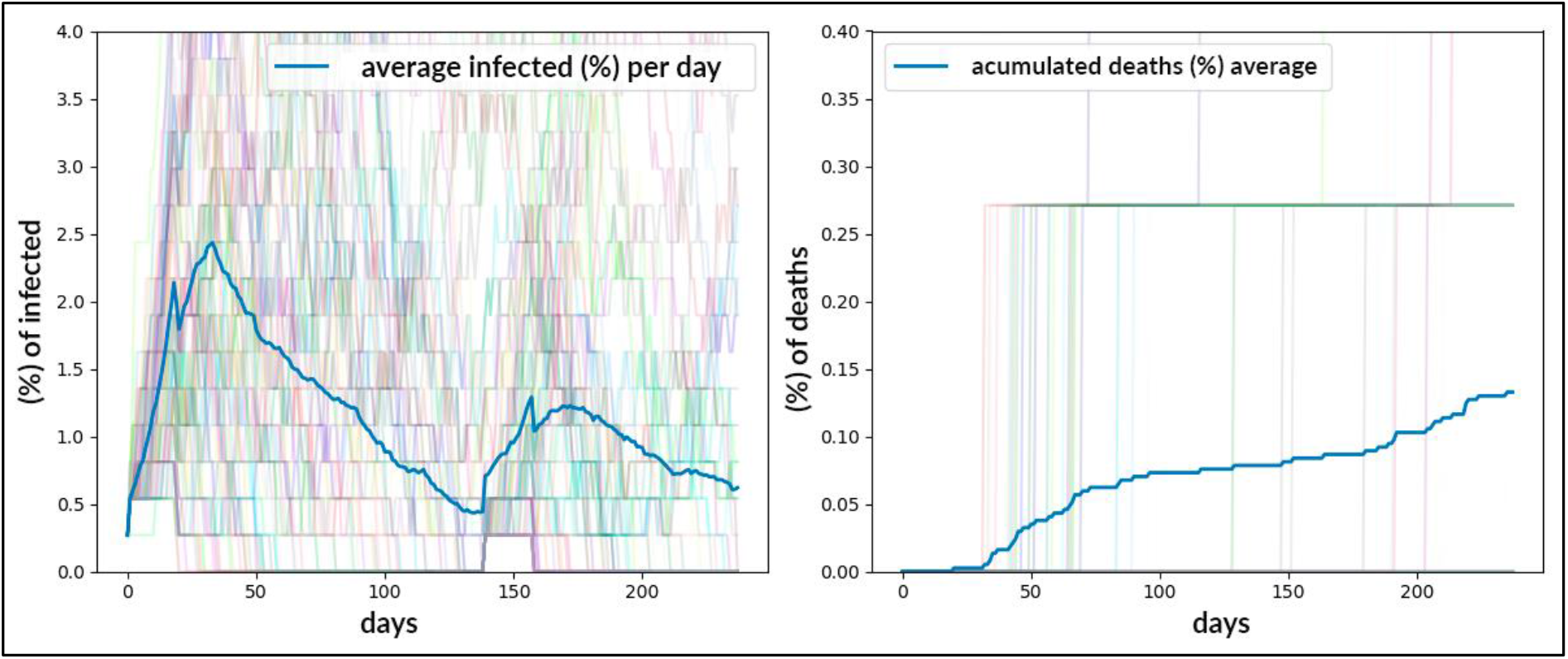
Dynamics of Virus Dispersion in the City of São Paulo after the reintroduction of an infected agent on the 138th day, using the official timeline (Table 1 in Appendix A) of social distancing, extended by 100 days at 40% isolation rate.

In all scenarios, even with a lower rate of social distancing, the maintenance of other types of NPI can help keep the transmission rate low, maintaining this protective scenario for longer and reinforcing the exhaustion of contagion networks for the city of São Paulo. As we discussed above, there are many studies showing the efficacy of social distancing, washing hands, and using protective masks to reduce the velocity of the virus’s spread (Leung 2020; Brauer, 2020; Bielecki, 2020; Cruz, 2020). The use of masks in particular (which became mandatory in the state of São Paulo as of May 7, 2020) has been shown to also reduce the intensity of COVID itself.

## Discussion and Conclusions

In this work we present preliminary studies of the dynamics of the coronavirus epidemic curve for the city of São Paulo using a simple and instrumental model. We intentionally worked with limited parameter values for the city of São Paulo to show that it is possible to reproduce the observed behavior of the epidemic without the need for ad hoc hypotheses like herd immunity in a population with a low infection rate.

Although models based on a deterministic SIR can be precise and complete when considering multiple variables that affect the virus’s spread (Alves, 2003; Mollison, 1995), this type of model (mean-field-like compartmental models) considers that an epidemic process evolves only when the density of susceptible individuals is greater than a limit value. In addition, this type of model is instrumental in evaluating scenarios but depends on ad hoc parameters (the transmission rate, for example) that are not experimental. Because of that, they are not as simple and intuitive as multi-agent models (Rahmandad, 2008).

The simplicity and the accessibility of our model allow us to show, for example, the effect of different territories’ vulnerabilities in the epidemic curve (Brauer, 2020; Winskill, 2020; Sumner, 2020). We showed that once the model is calibrated with data for a given region, we can make reasonable predictions (Ação Covid 19 a, 2020; Ação Covid 19 b, 2020; Ação Covid 19 c, 2020).

The simulations made using the MD Corona model indicate that at this moment (July 12, 2020) there should be a reduction in the number of cases and deaths in the Municipality of São Paulo, in line with the trend of a slight decrease in infections, deaths and beds occupied that was presented by official São Paulo state data (Seade, 2020)

This result is counterintuitive, as the opening of the economy, combined with a slight drop in the social-distancing index, could be expected to lead to an increase in the epidemic curve since the municipality is far from achieving so-called group (herd) immunity.

The model indicates that there are local “protection bubbles” against coronavirus infections. That is, although there are susceptible people, they are being isolated by a local barrier of immune people. A complementary explanation of this phenomenon is the theory of an exhaustion of the virus-contagion network, even after an initial worsening of the pandemic outbreak. This could be because social distancing, along with contagion-prevention practices (now widespread in society), may ensure a decrease in the speed of contagion or even the suppression of the virus in each sub-environment. In other words, this exhaustion is due to the formation of bubbles and of certain routines within the network of individuals maintaining social contacts among themselves.

In addition, the model reveals a kind of Urban Interiorization Dynamic of the virus, that is, local dynamics within the same territories generating new outbreaks of contamination (and immunization) in the same city. The virus is being passed from places and populations that have already had a certain amount of contact with the virus to other agents and places where that contact has not yet occurred. But the overall result can be a general drop in cases, deaths, and occupation of hospital beds.

Given the low number of immune people in the system, this reduction represents an unstable balance, which is fundamentally different from the expected stability of herd immunity. These protective bubbles can burst or the networks can be reinitiated, that is, new waves of transmission can occur if social distancing falls too much or there is a reintroduction of the virus in regions of the city where few agents have been infected and, therefore, have not become immune to the disease. But predicting these “burst bubbles” is extremely difficult without a policy of testing the population in the different districts of the Municipality.

These outbreaks can be mitigated by a policy of mass testing or selective testing and contact tracing. Only a few countries have managed to carry out the first strategy, such as Germany and South Korea, due to its high cost and the need for well-structured health services. This strategy is highly effective, and results in a proportional reduction in the number of cases when compared to the absence of this action. The selective-testing strategy is also reasonably effective in reducing the transmission chain as, in addition to testing severe hospitalized cases, it also tests for all suspected cases and those with whom they had contact, which, if positive for SARS-CoC-2, are isolated.

The municipality of São Paulo can mitigate the impact of transmission by implementing this strategy, as the city has a network of diagnostic RT-PCR tests and structured primary care services within a territorial base. But the effectiveness of this strategy also requires political and social factors that go beyond health policies. It requires: 1) mass communication and understanding by a large part of the population; 2) abandoning hopes of a “return to normality” through efforts to achieve “herd immunity” or by quickly developing an effective vaccine; and 3) social protection policies appropriate to existing social inequalities.

It’s important to stress that there are still several other possibilities that can be explored within this model, such as changing the average immunity time for the population (decreasing from one year to six months, for example), increasing or decreasing the average effective transmission probability of individuals, and increasing the number of people initially infected (or reintroducing more infected agents during the simulation).

Although the model is suitable for addressing heterogeneities within the city (differences in districts’ vulnerabilities, for instance), in our simulation we considered the city as a homogeneous unit and did not consider the heterogeneity within the city’s regions. To do that it would be necessary to build a more complex model with different interconnected environments. It would even be possible to incorporate information on urban mobility (urban and intra-urban displacements, etc.).

One important improvement could be made to the simulation analysis methodology. To deal with the stochastic behavior of the simulations, we presented the average curve for 100 identical scenarios. However, for the next studies we want to perform a more complete analysis, identifying the location of the behavioral-change thresholds. In order to do that, we have to analyze each simulation individually and then classify the different groups’ behavior.

These results could also be generalized to other cases, since this phenomenon is not exclusive to the city of São Paulo. The number of cases and deaths for the city of Manaus (Brazil) and Stockholm (Sweden) are also falling, despite the relaxation of social-isolation policies. To assert the generalization of this study we need to carry out further investigations.

## Data Availability

I declare that all data referred to in the manuscript is available

https://www.saopaulo.sp.gov.br/coronavirus/isolamento/

https://www.seade.gov.br/coronavirus/

---

**Appendix A.**
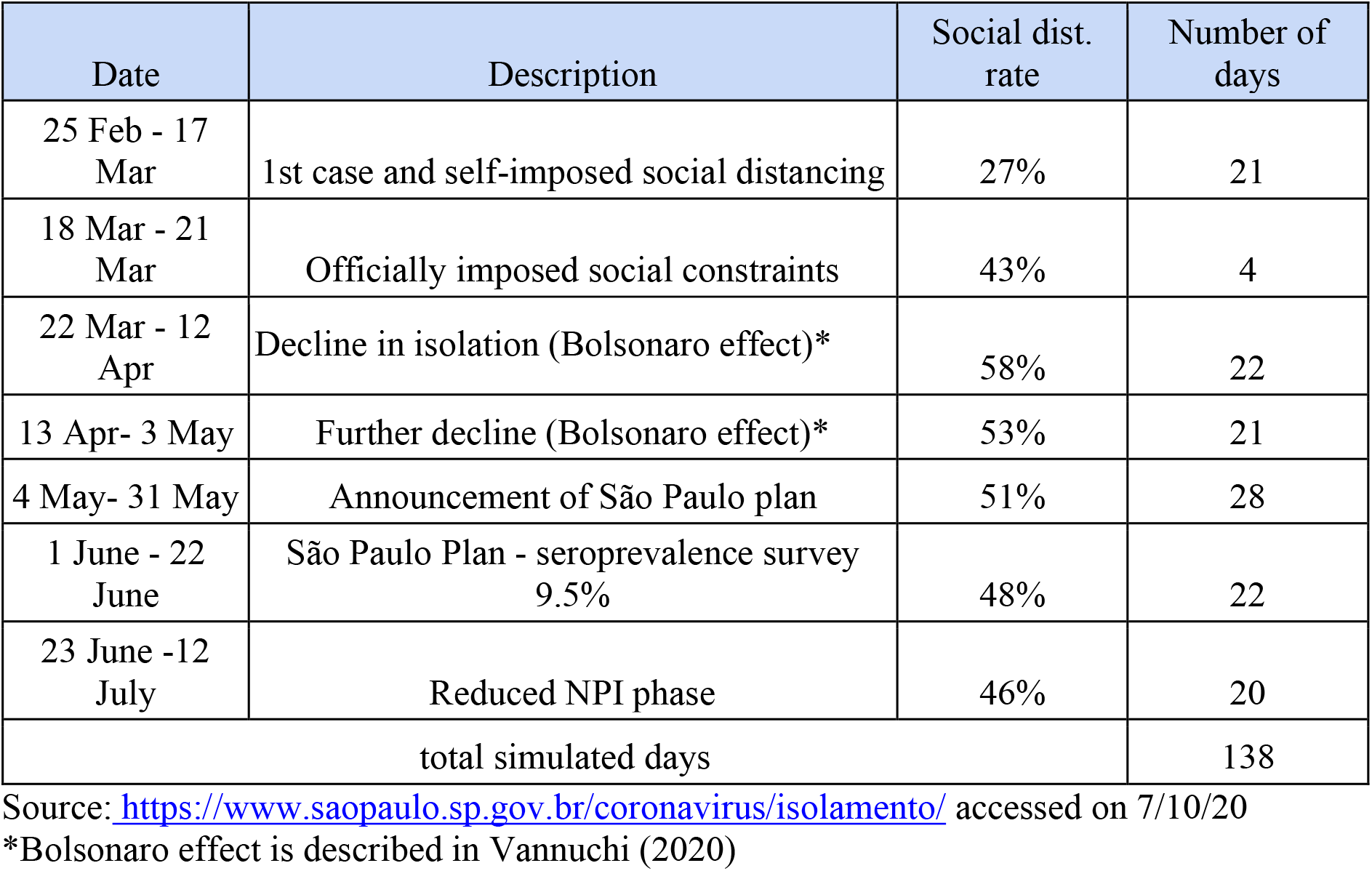
– Timeline of social distancing.

## Acknowledgements

We would like to thank the members of the Ação Covid-19 group for the support and fruitful discussions. This work was supported by Tide Setubal Foundation and the Federal University of ABC.

